# The effect of pre-anaesthetic assessment clinic: a systematic review of randomised and non-randomised prospective controlled studies

**DOI:** 10.1101/2021.06.04.21258364

**Authors:** Eirunn Wallevik Kristoffersen, Anne Opsal, Tor Tveit, Rigmor C Berg, Mariann Fossum

**Author notes:** Corresponding author: Eirunn Wallevik Kristoffersen, Department of Health and Nursing Science, University of Agder, Grimstad/Kristiansand, Norway.

## Abstract

**Objectives:** The aim of this systematic review was to examine the effectiveness of pre-anaesthetic assessment clinics (PACs) implemented to improve quality and patient safety in perioperative care.

**Design:** Systematic review.

**Data sources:** The electronic databases CINAHL Plus with Full Text (EBSCOhost), Medline, and Embase (OvidSP) were systematically searched from 1st April, 1996 to 4th February, 2021.

**Eligibility criteria:** The main inclusion criterion was that the study, using empirical quantitative methods, addressed the effectiveness of PACs.

**Data extraction and synthesis:** Titles, abstracts, and full texts were screened in duplicate by two authors. Risk of bias assessment, using the Joanna Briggs Institute critical appraisal checklist for quasi-experimental studies, and data extraction were performed by one author and checked by the other author. Results were synthesised narratively owing to the heterogeneity of the included studies.

**Results:** Seven prospective controlled studies were conducted. Most studies had a high risk of bias. Three studies reported a significant reduction in the length of the hospital stay, and two studies reported a significant reduction in cancellation of surgery for medical reasons when patients were seen in the PAC. In addition, the included studies presented mixed results regarding anxiety in patients.

**Conclusion:** This systematic review demonstrated a reduction in the length of hospital stay and cancellation of surgery when the patients had been assessed in the PAC. There is a need for high-quality prospective studies to gain a deeper understanding of the effectiveness of PACs.

**PROSPERO registration number:** CRD42019137724

**ARTICLE SUMMARY:** *Strengths and Limitations of this study:*

- Only prospective studies were included in this systematic review.
- The systematic review was conducted in accordance with international guidelines.
- Only seven studies were identified, highlighting the need for further research on pre-anaesthetic assessment clinics.
- Overall, the quality of the included studies was low, and the current practice possesses limited evidence base.

## INTRODUCTION

Anaesthesia constitutes an important part of surgery; however, it has the potential to activate physiological changes that can increase morbidity and mortality,[1] mainly depending on the patients’ preoperative health condition and age.[2] Hospitals are treating patients with complex, comorbid healthcare problems who undergo progressively extensive surgeries and interventions.[3,4] To ensure the quality and safety of anaesthesia and surgery, precise knowledge of the clinical characteristics of patients undergoing surgery is critical to the perioperative treatment plan.[2] Over the past 50 years, perioperative mortality, including anaesthesia-related mortality, has declined, with the most significant decline observed in developed countries,[1,5] mainly due to new anaesthetics, improved monitoring equipment and training, availability of recovery rooms, and improved airway management.[4] However, an Australian study reported that 14% of anaesthetic-surgical complications and 39% of deaths attributed to anaesthesia were associated with insufficient and/or inadequate preoperative evaluation.[6] A Danish retrospective investigation showed that the deaths among patients undergoing surgery could have been prevented by a thorough preoperative evaluation,[7] indicating that risk factors are both patient-and surgery-related and linked to organisational structures.[8] Future efforts should improve preoperative anaesthesia safety,[9] by improving planning and preparation for elective procedures and interventions.

In 1949, Lee discussed the value of the “anaesthetic outpatient clinic” in the preparation of patients for surgery.[10] Today, an increasing number of pre-anaesthesia assessment clinics (PACs) are supporting hospitals in handling the rise in the number and complexity of surgical procedures.[11] The PAC consultation, conducted by the anaesthesiologist, anaesthesia nurse, or both, is globally recognised as an evaluation method while optimising the patients’ medical condition prior to surgery and anaesthesia, and is considered essential in securing anaesthetic practice since it detects anaesthesia-related risk factors and high-risk patients, improves patient outcomes, prepares the patient physically and psychologically for anaesthesia, and ensures the patient’s most favourable condition for surgery and anaesthesia.[12-14] Considering the well-prepared patients and staff, several researchers posit that with PAC, the number of surgical cancellations, length of hospital stay, and mortality rate are reduced, and tests are minimised.[8,15,16] Others assert that patients feel less anxious regarding the subsequent anaesthetic and surgical processes and are highly satisfied with this service when PACs are used.[15,17,18]

As Turunen *et al*. state, research on PACs is scarce regarding costs, financial savings, the impact on patient safety and quality of care, accuracy of operative patients, and effect on preoperative nursing levels.[19] Survey results indicate that anaesthesiologists perceive day of surgery delays due to missing information as common, even with PAC consultations.[20] The present systematic review examines the outcomes of PAC as systematic work on quality and patient safety, including identifying the areas for improvement, implementing interventions, and ensuring that patient outcome improvement.

## METHODS

The aim of this systematic review was to examine the effectiveness of PACs in improving quality and patient safety in preoperative care. A further aim was to determine the gaps in existing knowledge for future research. Our systematic review followed the guidelines in the Cochrane Handbook for Systematic Reviews of Interventions [21] and was reported in accordance with the Preferred Reporting Items for Systematic Reviews and Meta-Analyses (PRISMA) guidelines.[22] The protocol was published in PROSPERO: CRD42019137724.[23] We had two review questions:

1. What are the effects of PACs on patient satisfaction, anxiety, and safety?
2. What are the effects of PACs on cancellation rate, cost, and efficiency?

### Search strategies

We performed a scoping search in different databases to identify the key terms for the literature search.[24,25] The final search was planned and conducted in close collaboration with a university librarian. On 11th September, 2018 we searched CINAHL Plus with Full Text (EBSCOhost), Medline, and Embase (OvidSP), and updated it on 4th February, 2021. Considering the lack of subject headings (e.g., MeSH) for PAC, we used text words such as preanaesthesia. The search in Medline is presented in Appendix 1. The search mode in CINAHL was Boolean/Phase, which supports Boolean searching or exact phrase searching. To ensure comprehensiveness, we used both the truncation and proximity operators. We limited the search to 1996 since this was the year one of the first known articles in this area was published.[23] Complementary methods to identify studies included following up on citations via Scopus, scanning the reference lists of relevant papers and included articles, and checking for relevant studies in clinical trials.[24]

### Eligibility criteria

Considering the aim of the review, the main inclusion criterion was that the study, using empirical quantitative methods, addressed the effectiveness of PACs. Specific study eligibility criteria were: (a) published in English or Scandinavian language, (b) scientific publication of original research, (c) reporting the outcomes of PAC, (d) PAC consultation with the patient present, (e) randomised or non-randomised prospective controlled studies, and (f) newly established PAC. We excluded: (a) editorials, discussion papers, and conference abstracts, (b) reviews, (c) instrument testing, (d) studies with children, and (e) retrospective studies.

### Study selection

All references identified in the search were transferred to EndNoteX9, where the duplicates were removed. Next, all unique references were transferred to the Covidence screening tool.[26] Study eligibility was ascertained independently by two authors, first at the title and abstract level, and subsequently at the full text. Inclusion was determined by consensus, and disagreements were resolved by consulting a third author.

### Quality assessment

We used design-specific checklists to assess the studies’ risk of bias. Given the methodological similarity of the included studies, only the Joanna Briggs Institute Critical appraisal checklist for quasi-experimental studies was used.[27] One author performed the risk of bias assessment, and the other checked the accuracy of the assessment.

Disagreements were resolved through discussion with a third author. Each of the nine checklist questions was answered no, yes, unclear (or not applicable).

### Data extraction and analysis

One author extracted data from each included study onto a pre-designed Excel spreadsheet, and another checked the extracted data for accuracy, consistency, and completeness. Extracted information included publication details, study design, setting, and characteristics of the patients, interventions, comparisons, and outcome (PICO). We requested information on the missing data; however, received no response from the author. If the PICO elements were sufficiently similar and statistical data were available, we had planned to conduct meta-analyses. However, the extracted data revealed substantial heterogeneity among the studies, and there were no randomised controlled trials (RCTs). Therefore, we performed a narrative synthesis, describing and comparing the main findings from the included studies, and discussing their methodological strengths and weaknesses.

## RESULTS

Figure 1. provides details of the study selection process. A total of 2250 records were identified in the first search and 742 in the second search. After removing duplicates, we screened 2372 records based on the title and abstract; of these, 179 records passed the full-text screening. We included seven studies that met the inclusion criteria.

**Figure 1:**
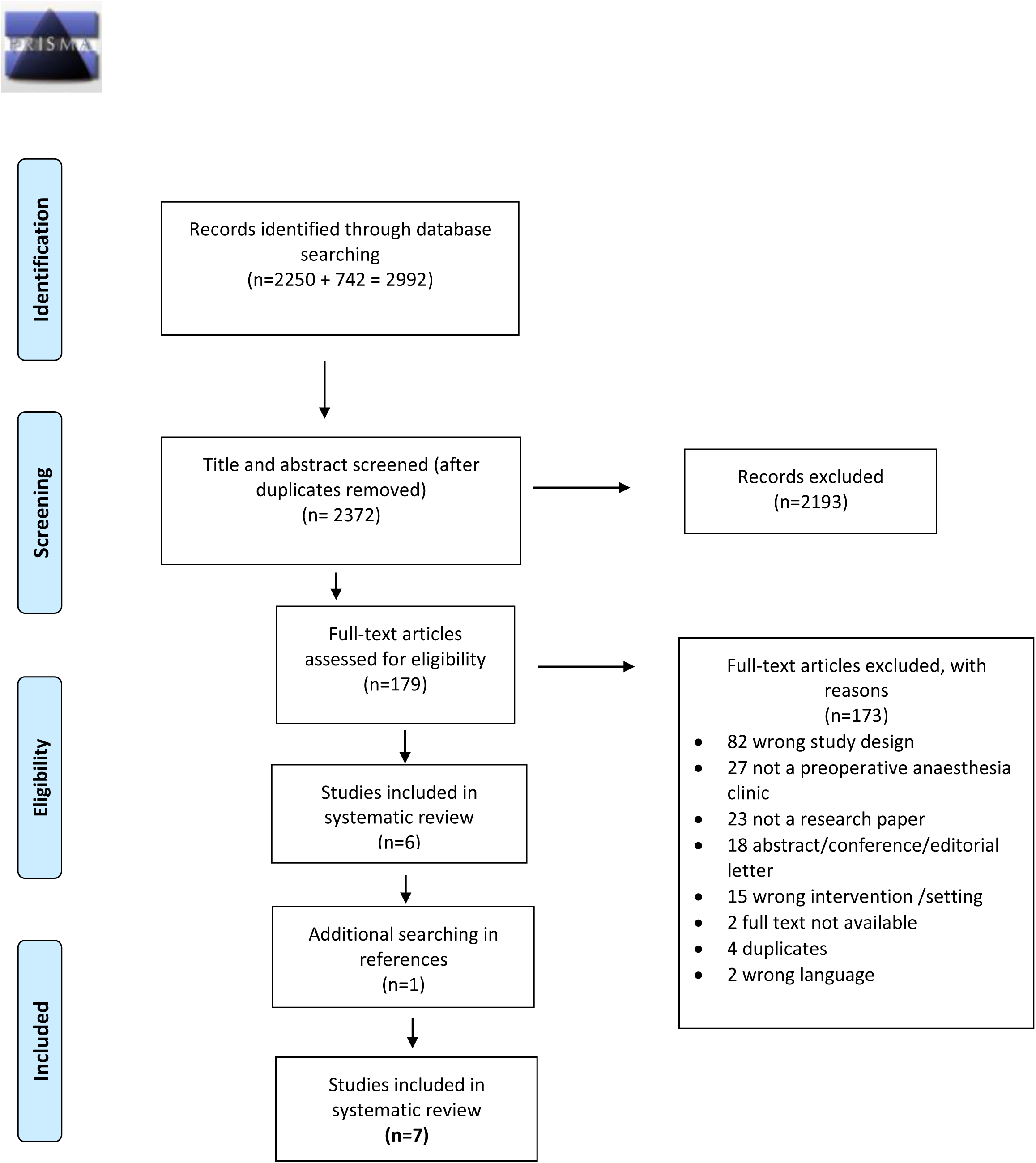
PRISMA Flow Diagram. *From:* Moher D, Liberati A, Tetzlaff J, Altman DG, The PRISMA Group (2009). *P*referred *R*eporting *I*tems for *S*ystematic Reviews and *M*eta-*A*nalyses: The PRISMA Statement. PLoS Med 6(7): e1000097. doi:10.1371/journal.pmed1000097

### Overall characteristics of the studies

The seven included studies are listed in Table 1. They were all in English and published in 2000–2017, with data collected in the years 1997–2015 (one did not provide this data collection information).[28] Based on our inclusion criteria, all were prospective controlled studies, but we found no RCTs. There was one controlled before-after study.[34] The other six studies had control groups but no baseline assessments, only assessments following PAC implementation. There were three 2-group non-parallel after-only studies,[29,30,32] and three 2-group parallel after-only studies [28], where one had a matched control group[31] and one had three follow-up assessments of one arm.[33] In total, the studies included 77411 patients.

**Table 1:** Description of included studies

| Author, Year, Country | Study design | Sampling time | Population | Intervention | Comparison | Outcomes |
| --- | --- | --- | --- | --- | --- | --- |
| Farasatkish, 2009[1]<br>Iran | 2-group after study | May 2007 through August 2007 | N=1716, open-heart surgery, ASA class III-IV | Pre-anaesthesia consultation clinic (3-10 days before surgery) | Usual care (within 24 h of surgery) | Cancellations |
| Kamal, 2011[2]<br>England | 2-group after study | April 2005 through April 2009 | N=1445, complex elective orthopaedic surgery, ASA class III-IV | Preoperative anaesthetic assessment clinic (timing not stated) | Usual care (day of surgery) | Admissions, length of stay, mortality, cost |
| Kamau, 2017[3]<br>Kenya | CBA | August 2000, April 2001, November 2001 | N=51, elective non-cardiac surgery, ASA class III | Pre-anaesthesia clinic consultation ($\geq 48$ h before surgery) | Usual care (day before surgery) | Anxiety (STAI score) |
| Klopfenstein, 2000[4]<br>Switzerland | 2-group after study (parallel) | No data | N=40, elective endoscopic urological surgery, ASA class I-III | Pre-anaesthetic consultation (1-2 weeks before surgery) | Usual care (the evening before surgery) | Anxiety (MAACL, VAS) |
| Lee, 2012[5]<br>China | 2-group after study (parallel) | March 2007 through November 2009 | N=352, elective surgery, ASA class I-IV | Anaesthesia consultation clinic ($\leq 3$ months before surgery) | Usual care (the evening before surgery) | Quality of recovery score), cost, cancellations, length of stay, satisfaction, anxiety (VAS), willingness to pay (WTP) |
| Mendes, | 2-group | April 2007 | N=52254, | Preoperative | Usual care (timing | Cancellations, |
| 2005[6]<br>Brazil | after study<br>(parallel) | through<br>June 2007 | surgery, ASA<br>class not stated | outpatient evaluation<br>clinic (timing not<br>stated) | not stated) | length of stay |
| van Klei,<br>2002[7]<br>The<br>Netherlands | 2-group<br>after study | November<br>2012 | N=21553,<br>elective surgery,<br>ASA class mainly<br>I-II | Preoperative<br>outpatient evaluation<br>clinic (average 3 weeks<br>before surgery) | Usual care (day<br>before surgery) | Cancellations, same-<br>day admissions,<br>length of stay |

ASA: American Society of Anesthesiology; CBA: controlled before-after; MAACL: Multiple Affect Adjective Check List; STAI: State-Trait Anxiety Inventory; VAS: visual analogue scale; WTP: willingness to pay

Considering the intervention, PACs in all studies consisted of an outpatient service whereby patients were checked for medical conditions that are important for anaesthesia and informed regarding what to expect on the day of surgery. However, the terminology used for PACs varied; they served different surgical specialities, and the pre-anaesthesia consultation was conducted from ≥48 h to ≤3 months before the surgery. Three were implemented in a university hospital,[31,33,34] one in a teaching hospital,[30] one in a medical centre,[32] and one in a general hospital[29] (one study did not specify the context).[28] The person conducting the pre-anaesthesia consultation also varied: in five studies, it was the anaesthesiologists,[28-31,33] in the other studies it was (also) the orthopaedic senior house officer,[29] the consultant or resident,[34] or the physician.[32] In three studies, nurses were part of the team assessing the patients.[29-31] The comparison group in all studies was usual care, which generally involved performing a preoperative anaesthetic evaluation the day before the surgery on the admitted patients.

Of the 77411 patients in the studies, 9626 and 15531 patients were in the intervention and control groups, respectively. One study did not specify the number of patients in the intervention and control groups, but only the total number of surgeries performed.[33] Five studies reported data for sex, showing that 51% of the patients were women and 49% were men (12129 vs. 11583).[28,30-32,34] There were more females than males both in the intervention (4345 vs. 4134) and the control groups (7784 vs. 7449). Five studies reported data for age showing that all the patients were over 20 years old and grouped within the American Society of Anesthesiology (ASA) category.[28,30-32,34]

The patients were scheduled to undergo a variety of surgeries, including orthopaedic,[29-31,34] urology,[28,30,31,34] general,[30,31,34] heart,[32] gynaecology/obstetrics, [30,31,34] vascular surgery,[30] ophthalmology,[30] maxillofacial/dental surgery, [30,34]neurological surgery,[30] and one did not specify the type of surgery.[33] In five studies, the type of anaesthesia was not specified,[29,30,32-34] and two studies reported patients for general and/or regional supplement.[28,31]

The patients included had previous anaesthetic experience in one study,[28] previous and no previous anaesthetic experience in another,[34] and five studies did not report this data.[29-33] Limited background characteristics of the patients were reported in two studies.[29,33] One stated that the patients included had ASA 3 or 4 and a body mass index of more than 40. However, no ASA number, sex, or age was reported in the article.[29] Mendes *et al*. did not report any background characteristics of the included patients.[33]

### Description of the studies’ risk of bias

Figure 2. shows the results of the risk of bias assessment. In all seven included studies, the cause and effect were clear. The majority of the studies measured outcomes in the same way and used appropriate statistical analyses. Several studies had limitations of follow-up and similarity in care and participants. None of the patients had multiple pre-and post-measurements.

**Figure 2.**
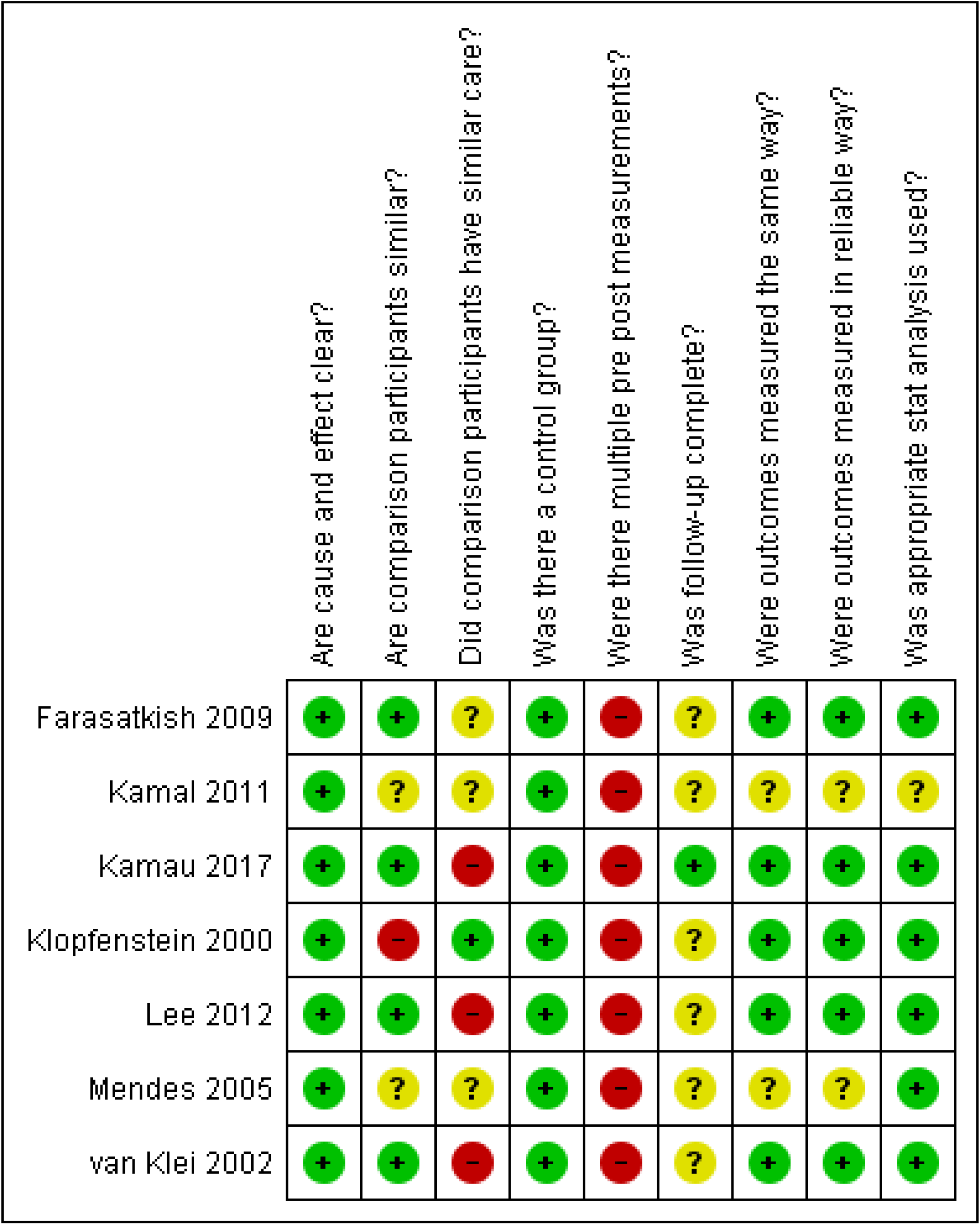

### Outcomes of the included studies

The outcomes of the included studies are described separately below.

#### Satisfaction

One study reported satisfaction as an outcome.[31] The summarised patient satisfaction with the anaesthetic consultation score out of 100 showed that patients in the PAC group were more satisfied (mean difference, 2.10%; 95% confidence interval [CI], 0.51–3.70%; *p*=0.01).[31] There was no statistically significant difference between the two groups in the mean patient satisfaction with perioperative anaesthesia care score out of 5 after surgery (mean difference 0.01%, *p*=0.94).[31] The mean quality of recovery (QoR) score (range, 0– 18) following anaesthesia on the first day of surgery was similar between the intervention (13.17±2.73) and control (13.31±2.65) groups (*p*=0.67).[31] The QoR measure is the patients’ health-related quality of life.[35]

#### Anxiety

Three studies reported anxiety.[28,31,34] Two studies reported the visual analogue scale (VAS), one rated from zero (no anxiety) to ten (very high anxiety),[28] another used a 100 mm horizontal line with “not anxious at all” to “extremely anxious”[31] In one study, the median VAS anxiety score was 3 (0–5) in the intervention group and 5 (2–8) in the control group (*p*=0.0038).[28] In another study, there were no significant differences between the control and intervention groups for levels of anxiety (VAS), surgery (26 vs. 25, respectively, *p*=0.12), and anaesthesia (20 vs. 19, respectively, *p*=0.60).[31] The median Multiple Affect Adjective Check List (MAACL) score with possible range scores from 0 to 21 (higher scores indicating greater levels of anxiety) was 3 (0–9) in the intervention group and 6.5 (2–12) in the control group (*p*=0.0053).[28] The differences in the State-Trait Anxiety Index (STAI) score, which is composed of 40 questions rated on a 4-point Likert scale, was 1.51, 95% CI: 1.02–2.02%, *p*=0.0051).[34] The results on anxiety in these two studies were significant. However, Kamau *et al*.[34] found no differences when they examined anxiety and the influences of sex, duration of hospital stay, and prior anaesthesia experience.

#### Mortality

One study reported the mortality rates.[29] Patients attending the High Dependency Unit (HDU), Intensive Care Unit (ICU), and Post-anaesthesia Care Unit (PACU) following complex orthopaedic surgery had a significant reduction in mortality rate after being assessed in the PAC, from 18 (6.1%) of 298 patients before to 14 (1.2%) of 1147 patients after *p*=0.001.[29]

#### Cancellation rate

Three studies reported a reduced cancellation rate following the establishment of a PAC.[30,32,33] One of the included studies had 316 (2.0%) cancellations for medical reasons before the introduction of PAC, and 79 (0.9%) after, and a difference of 1.02% (95% CI, 0.31– 1.31%). After adjustment, the odds ratio was 0.7 (95% CI, 0.5–0.9%).[30] The overall cancellation of surgery was reduced from 1027 (6.3%) to 393 (4.6%) following surgery, and a difference of 0.9% (95% CI, 0.3–1.0%) when patients were assessed in PAC.[30] Mendes *et al*.[33] found a decrease in overall cancellations from year 1 (39.3%) to year 4 (15.9%), *p*≤ 0.05. There were 469 (number of cancellations)/10639 (number of surgeries performed) due to medical reasons in the first year of this study. The following year, a considerable increase above the baseline values in the intervention group was observed, followed by a progressive decrease in the last year with 391 (number of cancellations)/10397 (number of surgeries performed).[33] Farasatkish *et al*. reported that of the 1716 patients studied, 15.1 % of cases cancelled in the two groups. The cancellation rates in the control group were 146/866 (16.8%), and the cancellation rate in the intervention group was 113/850 (13.29%) p=0.046. The most common reason for cancellation was incomplete medical work-up 51/146 (35%) in the control group and 32/113 (28%)in the intervention group, p=0.03).[32] Lee *et al*. found similar rates for surgery being cancelled on the scheduled date for the intervention group compared to the control group (2.3% vs. 3.4%, *p*=0.75).[31]

#### Costs and willingness to pay

Two studies reported the costs.[29,31] One study reported a total saving of £ 486.62 per patient after establishing a PAC.[29] Another study reported a significantly lower preoperative cost per patient in the intervention group compared to the control group (mean difference, $ 463; 95% CI, -$648 to -$278 per patient, *p*<0.01).[31] However, the mean difference in the total perioperative treatment cost was not significant, even after adjusting for cancellation on the day of surgery costs.[31] The intervention group patients were willing to pay (WTP) significantly more than the median WTP (US $13) for a clinic consultation at the PAC than the control group.[31]

#### Length of stay

The length of stay was reported in four studies.[29-31,33] Mendes *et al*.[33] found a significant decrease in mean hospital stay for patients from 6.2 to 5.0 days (p ≤ 0.001) during the four years of this study. Van Klein *et al*.[30] found that the total admission time significantly decreased from a mean of 8.8 days (before) and a mean of 8.1 days (after) and 0.92 (0.90–0.94). After adjusting for age, sex, and introduction date of PAC this difference was 0.92 (0.90–0.94).[30] Kamal *et al*.[29] found a significant reduction in the length of stay in the high dependency unit from 2.1 days to 1.6 d (*p*=0.01), and in the intensive care unit from 2.3 days to 1.9 days (*p*=0.01). In the last study, no significant changes were found in the median duration of postoperative stay between the intervention and control groups.[31]

#### Organisation planning and efficiency

Organisation planning and efficiency have been reported in two studies.[29,33] One study found statistically significant changes in the reduction of unplanned admissions to the PACU (65/298 [22%], 111/1147 [10%], *p*=0.001), ICU (4/298 [1.3%], 4/1147 [0.4%], *p*=0.01), and HDU (4/298 [1.34%], 20/1147 [1.7%], *p*=0.01) after implementing a PAC.[29] The planned admissions in the ICU (4/298 [1.3%], 18/1147 [1.6%], *p*=0.01), and HDU (14/298 [4.7%], 85/1147 [7.4%], *p*=0.1) increased after implementing a PAC.[29] The number of PAC evaluations increased from year 1, 4704 to year 4, 13990 (*p*≤ 0.001).[33] The number of outpatient procedures increased from 2170 (year 1) to 1943 (year 4) (*p*≤0.001), and the inpatient procedures decreased from 9556 (year 1) to 8449 (year 4), (*p*≤ 0.001).[33]

## DISCUSSION

This systematic review summarises the effectiveness of PACs in improving quality and patient safety in general hospitals and determines the gaps in existing knowledge for future research. Seven studies that met the inclusion criteria were included. We present the main results and infer the implications for research and practice in the following text.

Cancellation on the day of surgery has undesirable effects on both the patients and the hospital system.[14] Thus, studies have found that late patient-related cancellations could totally or partially be prevented,[36] if they were addressed during preoperative evaluations.[14,15] This is confirmed by several studies in this systematic review that found a reduction in surgery cancellation after implementing a PAC.[30,32,33] However, Lee *et al*. found no significant changes between the intervention and control groups.[31] Mendes and colleagues found that the number of cancellations for medical reasons after PAC implementation decreased in the first year of implementation. In the second and third years, they were higher before the number dropped to below baseline.[33] These conflicting findings might show that hospitals operate in a specific context, with unique populations, processes, and microsystems, which may encounter unique obstacles making implementation difficult. Patient-focused interventions need to consider barriers, facilitators, and interrelationships between systems, staff, and interventions to increase the likelihood of sustainable success.[37] In addition, Kamau *et al*. also indicated that PACs lead to more planned admissions to the ICU, HDU, and PACU, which is more predictable for patients, staff, and administrations.[34]

Another main finding of this systematic review was a significant reduction in the length of hospital stay following patients’ examination in a PAC; however, a small number of studies with low quality were considered. Nevertheless, similar results were found in another systematic review claiming that perioperative systems support hospitals to address the expected growth in the number and complexity of surgical procedures being performed.[15] However, Lee *et al*. indicated that the reason for the reduced length of hospital stay was the mean duration of stay before surgery in the intervention group.[31] This indicates that when patients are examined in the PAC and well prepared with information, consultations, and tests, they do not need to be hospitalised until the day of surgery. A survey focusing on patients operated showed that if they had a choice, 75% do not wish to be admitted to the hospital until the same day of operation. One of the main reasons was to spend less time in the hospital.[38] However, an updated systematic review on the effectiveness of nurse-led preoperative assessment services for elective surgery found that the included articles had a reduced length of stay. The included studies had low methodological quality, and therefore, the authors could not conclude that this service leads to reduced length of hospital stay.[16]

The evidence from this systematic review is insufficient to conclude whether patients have reduced anxiety when assessed using PAC. The included studies used different instruments to measure the levels of anxiety, and the results could not be pooled. In addition, previous studies have shown that anxiety levels were higher in women.[39] Seventy-eight per cent of the participants were women in one of the included studies in this systematic review and might result in a bias in this study.[34] Anxiety was also statistically higher in patients who underwent general anaesthesia than in those who underwent regional anaesthesia.[40] The included studies on anxiety included both patients with general and regional anaesthesia, which might also be biased. Furthermore, the patients included in this review had both former surgical experience and no experience with surgery. However, studies have shown that former experience with anaesthesia and surgery reduces the risk of preoperative anxiety.[41]

Assessment of PAC was significantly associated with reduced mortality following complex orthopaedic surgery.[29] Previously published retrospective studies found similar results, but with other types of surgery.[42,43] A Danish study found that deaths attributed to anaesthesia were associated with insufficient or inadequate preoperative evaluation.[7] Furthermore, a previous study pointed out that the risk factors are not only patient-related but also organisation-related,[8] and that some hospitals have perioperative care and teams that are better at identifying and rescuing perioperative complications.[44,45] However, Blitz *et al*. argued that PAC should focus on early patient engagement strategies, interdisciplinary team communication, detailed perioperative care plans, and patient documentation in the electronic health record. This record should be open for review by the perioperative team to preserve patient information and safety. The value of a PAC lies in its ability to improve the quality of the perioperative process by designing a more robust system for preoperative assessment and preparation.[42] The importance of safety in anaesthesia is a vital component in anaesthesia practice, and the use of PACs contributes to this critical area.

### Strengths and limitations of the study

Most review steps were performed in duplicate or independently by two researchers, and agreement was reached in a consensus meeting. However, grey literature, such as government and institutional documents, was not included and might be a limitation to this study. Since countries have different organisational structures in their healthcare systems, we did not set inclusion criteria concerning who performed the patient’s preoperative assessment. However, the European Society of Anaesthesiology guidelines recommend that anaesthesiologists complete the preoperative assessment, while trained nurses or anaesthesia trainees perform the screening.[8] A preoperative evaluation performed by an internist has been associated with increased length of stay and increased postoperative mortality.[46] This systematic review’s results were possibly affected by the heterogeneity in the types of staff performing the preoperative assessment.

We opted to include only the studies with the highest internal validity. Thus, we excluded several retrospective studies. Nonetheless, the remaining studies’ risk of bias was fairly high, and they were heterogeneous. As a result, meta-analyses were not statistically appropriate.[25] The included studies’ designs could not rule out selection bias and confounding, and the strength of the evidence should be assessed cautiously. Many studies did not make adjustments for several confounders, which could be responsible for the observed effects. Several studies lacked descriptions of the methods used and the patients included, which lowered the transparency. It is not very reassuring that many such studies were unable to deliver more thorough evidence to guide practice and should be assessed cautiously. The results are relevant to health care services, which should focus on the well-being and safety of the patients.

### Implications for future research and practice

This systematic review identified the ambiguity in the PAC interventions offered to the intervention group. In many studies, it was evident that the methods used in these studies were not always clearly described, and high-quality research is needed in this field. The included studies in this review did not contain any results of reduced preoperative tests, such as blood tests, on patients before surgery when patients attended the PAC,[47,48] and earlier surgical room entry time for patients assessed in PACs,[49,50] similar to previous retrospective studies. Other implications for future research might be the organisation structure of different PACS and their functioning. The use of technology, such as streaming services, facilitates different types of patient groups and might be more important with the appearance of Covid-19 in reducing human contact and spread of the virus.

## CONCLUSION

This systematic review suggests that PAC use reduces the length of hospital stay, and the majority of the studies had reduced the cancellation rate in hospitals. These findings are an essential contribution to the current evidence in this field. In addition to further research in this field, the demand for increased high-quality studies to capture robust data describing the quality of care and clinical outcomes for patients requiring anaesthesia. This step demands increased focus and funding for this specific area of health services research and could, therefore, lead to new implementations of PAC`s in health care services and further develop patient safety in perioperative care.

## Supporting information

Medline search

## Data Availability

All data relevant to the study are included within the article or have been uploaded within supplemental files.

## Acknowledgement

We thank the librarian Ellen Sejerstedt, University of Agder, Kristiansand, for helping with the search strategy and removal of duplicates.

## Author statement

EWK, MF, AO and TOT: Study design

EWK, MF: Search and screening of the articles

EWK: Data extraction

MF, AO, RB and TOT: Control of data extraction

EWK, MF, AO and RB: Quality assessment of the included articles

EWK: Drafting of the manuscript

MF, AO, RB, and TOT: Contribution to and review of the final version of the manuscript

MF, AO, RB and TOT: Supervised the study

