## Supplementary material for "The effect of pre-anaesthetic assessment clinic: a systematic review of randomised and non-randomised prospective controlled studies": Medline search

Database: Ovid MEDLINE(R) ALL <1946 to February 03, 2020>
Search Strategy:

--------------------------------------------------------------------------------

1 ((pre-operativ* or preoperativ* or pre operativ*) and (assessment* or measurement* or evaluat*)).ti. (6182)

2 (clinic* or unit* or nurs* or outpatient* or ward* or center* or centre*).ti. (1472922)

3 1 and 2 (478)

4 ((preanaesthe* or pre-anaesthe* or pre anaesthe* or pre-anesthe* or preanesthe* or pre anesthe*) adj4 (assessment* or measurement* or evaluat* or clinic* or nurs* or unit* or outpatient* or ward* or center* or centre*)).ti,ab. (572)

5 (((pre-admiss* or preadmiss*) adj4 (assessment* or measurement* or evaluat* or clinic* or unit* or nurs* or outpatient* or ward* or centre* or center*)) and (surg* or anaesthe* or anesthe* or preoperativ* or pre-operativ* or preanaesthe* or pre-anaesthe* or pre anaesthe* or pre-anesthe* or preanesthe* or pre anesthe*)).ti,ab. (241)

6 ((anaesthe* or anasthe* or anesthe*) adj4 outpatient* adj4 clinic*).ti,ab. (138)

7 ((pre-admiss* or preadmiss*) adj4 (center* or centre*) adj4 (evaluat* or assessment* or measurement*)).ti,ab. (4)

8 or/3-7 (1409)

9 limit 8 to yr="1996 -Current" (1105)

***************************
